# Economic evaluation of a hospital-initiated tobacco dependence treatment service

**DOI:** 10.1101/2025.03.21.25324390

**Authors:** John Robins, Gary Alltimes, Ann McNeill, Irem Patel, John Moxham, Stephanie Duckworth Porras, Arran Woodhouse, Andrew Stock, Debbie Robson

## Abstract

The treatment of tobacco dependence in patients admitted to hospital is a priority for the National Health Service in England. We conducted an economic analysis of a pilot intervention adapted from the Ottawa Model of Smoking Cessation, implemented in a major teaching hospital in London, England.

The cost-per-patient, cost-per-quit and Incremental Cost Effectiveness Ratio were estimated for 673 patients who smoked and who received the intervention after being admitted to one of 11 acute wards between July 2020 and June 2021. Patient-level readmission costs and bed-days from six months after discharge were compared between the intervention group and a group of benchmark patients who smoked and who did not receive the intervention.

The total cost of the intervention was £178,105. On the basis of 104 patients who reported not smoking at six months, the cost-per-quit was £1712.55. Among 611 patients who were successfully matched to a benchmark cohort, re-admissions for patients in the intervention group cost £492k less than their benchmark equivalents over 21 months from January 2021 to September 2022 (£266k vs £758k), incurred 414 fewer bed days (303 vs 717), and re-admitted at a lower rate (5% vs 11%). Lower readmission rates and costs were associated with the intervention regardless of patient smoking status at six months, except among those who had opted out.

A pilot tobacco dependence treatment intervention implemented in an acute hospital setting in London demonstrated value for money through reduced readmission rates and costs among all patients who received it.

## 1 Background

Tobacco smoking remains the modifiable mortality risk factor that accounts for more years of life lost than any other (1). It increases the risk of a multitude of diseases, especially lung cancer, chronic obstructive pulmonary disease (COPD) and coronary heart disease (CHD), and is the largest avoidable cause of disability and social inequalities in health in the UK (2). In 2019/20 there were an estimated 506,100 admissions to hospitals in England that were attributable to smoking, costing an estimated £850m (3).

Recently, there has been significant new investment within the National Health Service (NHS) in England that focuses on the treatment of tobacco dependency for patients admitted to acute medical care, to psychiatric hospitals, and to maternity services (4). The NHS Long Term Plan committed to ensuring that all patients admitted to hospital who smoked would be offered NHS-funded tobacco dependency interventions by 2024. Recommended interventions included the Ottawa Model for Smoking Cessation (OMSC) - an “opt-out” model which incorporates the systematic identification of the smoking status of all admitted patients, followed by brief advice, personalised bedside counselling, timely nicotine replacement therapy (NRT) and/or pharmacotherapy, and follow-up after discharge (5). Hospital-initiated smoking cessation interventions have been found to be effective in increasing quit attempts and quit success (5–10) and reducing readmission rates and mortality (5,11).

There is less evidence regarding the cost-effectiveness of hospital-initiated smoking cessation interventions, although where such research has been conducted the results are favourable, despite differences in delivery of the intervention and the methods of evaluation (12). A 6-month pilot of the CURE Project (Conversation, Understand, Replace, Experts and Evidence-based interventions)—an opt-out model similar to the OMSC—in Manchester, England, estimated a cost per quit of £475, based on the 22% quit rate of smokers followed-up for 12 weeks after discharge (10,13). The estimated gross financial return on investment was £2.12 per £1 invested over a payback period of 4 years. When using metrics which incorporate the value of improved patient health among those who quit smoking, the public value return on investment was £30.49 per £1 invested, and the cost per Quality-Adjusted Life Year (QALY) was £487 (13).

### 1.1 Current study

In order to support the continued funding of OMSC-type interventions, further evidence is required to demonstrate effectiveness, both in terms of patient outcomes and cost. This study estimated the cost per patient and cost per quit of an adapted OMSC intervention, implemented in an acute hospital in Southeast London, England. Additionally, we extended previous economic evaluations by using patient-level readmission data from a matched cohort of patients who smoked but did not receive the intervention to estimate the variance in readmission costs and bed-days associated with the intervention.

## 2 Methods

### 2.1 Setting

The study used data from 673 patients who smoked, admitted to one of 11 acute medical and surgical wards in a large teaching hospital run by King’s College Hospital NHS Foundation Trust in Southeast London, between 1st July 2020 and 30th June 2021 (herein referred to as the ‘OMSC group’). Median length of stay was six days. The 673 patients comprise all of the patients seen and assessed by the Tobacco Dependence Specialists (TDS) during their hospital stay, representing 40.2% of all identified smokers admitted to the 11 wards during the same period, and 6.2% of all admissions . Post-discharge phone calls were made to all 673 patients at 30-day intervals up to six months, except where a patient had specifically requested to opt-out. Clinical audit approval was obtained from the NHS Trust prior to the study. Further details of the intervention and patient-related outcomes from a larger study involving this and another hospital Trust are described in Robins et al. (2024).

### 2.2 Economic evaluation

#### 2.1.1 Cost per quit

The total costs of running the adapted OMSC intervention were calculated and applied to the number of patients reporting being a non-smoker at 180 days after discharge. The Incremental Cost Effectiveness Ratio (ICER)—the incremental cost per life year gained associated with the intervention—was calculated using the formula and tables provided in Stapleton & West (2012).

#### 2.2.2 Patient-level costs

Patient-level costs were estimated using the NHS Trust’s Patient-Level Information and Costing Systems (PLICS) data, which combines activity, financial, and operational data to cost individual episodes of patient care (15). The PLICS model calculates patients’ resource usage, overhead, and indirect costs to derive the total cost of each patient recorded by the Trust’s Information department. The data from the OMSC group were matched to the Trust’s PLICS to retrieve activity and cost data. Comparing patient ID and the admission and discharge dates from the OMSC group to PLICS returned 653 costed patient spells, with 20 patients unmatched and discounted from the remaining calculation process. Patients who died within the year following discharge (n=22), or who had incomplete follow-up data (n=20), were also excluded from the analysis, leaving an analytic sample of 611 OMSC group patients.

#### 2.2.3 Benchmarking

The benchmarking focussed on the frequency, cost, and duration of patient re-admissions, by comparing patients in the OMSC group to similar patients that had not received the adapted OMSC intervention, i.e. patients who smoked but who had not been seen by a TDS.

The benchmark patients were drawn from inpatient admissions at the Trust between July 2020 and June 2021, after extreme outliers in terms of length of stay had been removed. Through data collected on admission, we were able to identify patients that had declared themselves as smokers. This dataset was then matched against the OMSC group data to find an appropriate benchmark cohort for each OMSC patient. See **Figure 1** for summary of benchmarking process.

**Figure 1.**
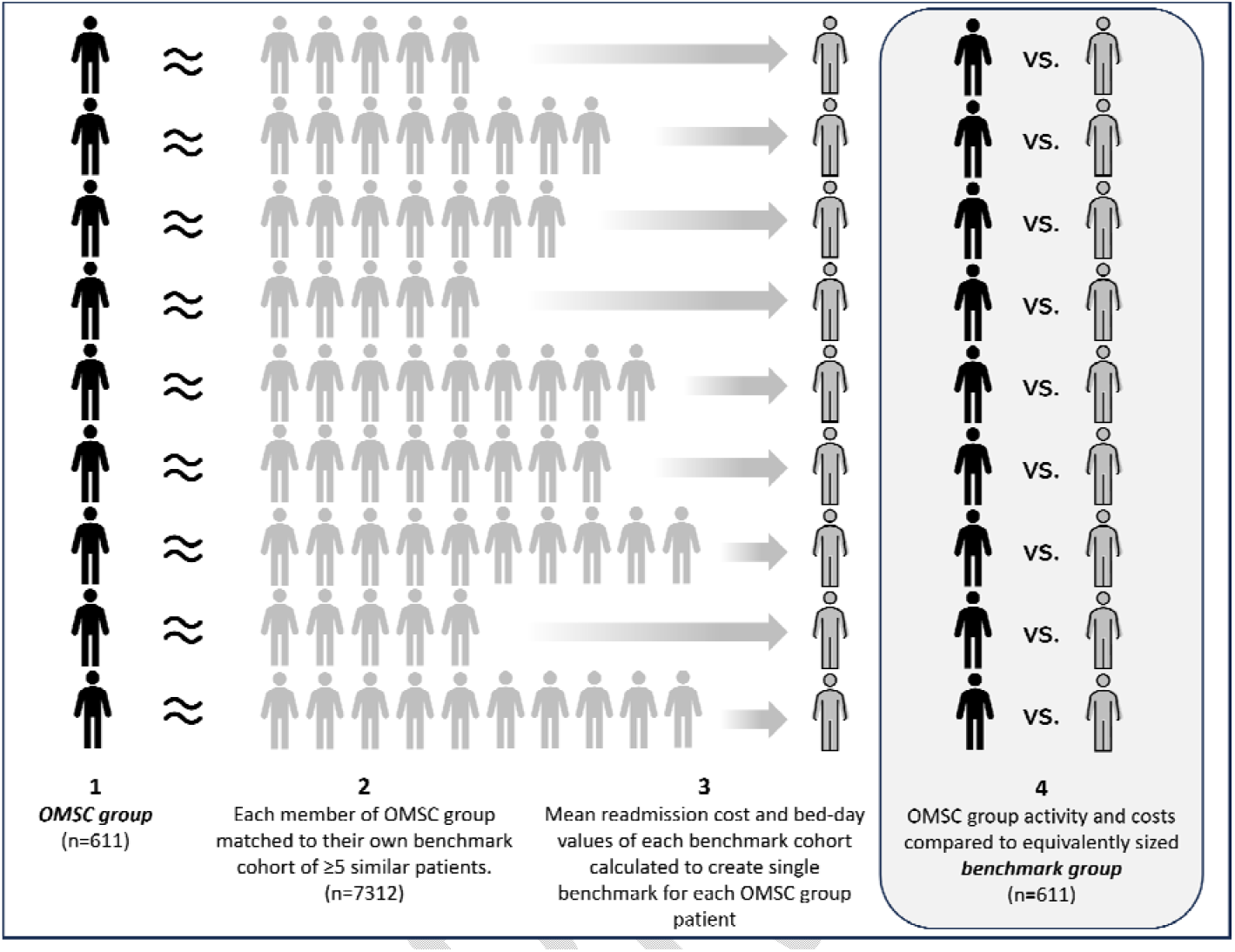
Benchmarking process: (1) After excluding patients with incomplete follow-up data or who could not be matched on PLICS, each patient in the OMSC group was matched to their own benchmark cohort, (2) consisting of at least 5 patients who did not receive the OMSC intervention but who were sufficiently similar in terms of demographic and clinical characteristics (i.e. smoking status, how many smoked per day, Index of Multiple Deprivation decile, age, delivery [elective/emergency], Healthcare Resource Group root code and sub-chapter). (3) From each matched benchmark cohort, the mean readmission cost and bed-day values were calculated and used to represent the benchmark to which the readmission costs and bed-days of the OMSC patients were compared (4). Collectively, these mean values form the “benchmark group”.

##### 2.2.3.1 Matching rules & loops

The OMSC patients were compared to the benchmark patients using combinations of the following criteria to ensure appropriate comparisons: how many cigarettes smoked per day, point of delivery (elective/emergency), Healthcare Resource Group (HRG) root code and sub-chapter (i.e. the clinical classification of patient events that have been judged to consume a similar level of resource), Index of Multiple Deprivation (IMD) decile, and ten-year age range (selected on the basis of being broad enough to return a benchmark cohort but without being too broad as to make the application of age-related criteria meaningless).

Additionally, a rule was set that a minimum of 5 comparable patients must be found in each benchmark cohort, to reduce the likelihood of the results being skewed by any one benchmark patient. These criteria were applied in 14 matching loops against the OMSC group to find the most appropriate benchmark cohort for each OMSC patient. **Supplementary Table S1** shows the matching process noting where each criterion has been matched or dropped per loop, as well as the number and proportion of OMSC group patients matched to cohorts in each loop.

##### 2.2.3.2 Re-admissions calculation

PLICS data was assessed for all patients across both cohorts to identify any re-admissions after 180 days from original discharge from the Trust. The 180-day cut-off was chosen to ensure re-admissions would be adjacent or subsequent to the final follow-up phone call to patients in the OMSC group, in which they would have reported their smoking status six months after discharge. It also focuses on a period when a patient’s reduction in smoking would be more likely to have a tangible effect on their health and likelihood of re-admission (16). The PLICS data period for assessing re-admissions covered the period from 1^st^ January 2021 to 30^th^ September 2022.

Readmission rates in the OMSC group were compared to those of the benchmark group, both overall and stratified by the self-reported smoking status at 6 months in the OMSC patients (i.e. non-smoker, smoker, unknown, or opted out). Comparisons by 6-month smoking status were only possible on the basis of the OMSC group smoking statuses, as the post-discharge smoking statuses of the benchmark group were unknown. Total readmission bed days and total readmission costs were calculated for the OMSC and benchmark groups, with the latter being derived using the mean values of each OMSC patient’s matched benchmark cohort (**Figure 1**). Bed days and costs per-patient-readmitted were calculated for the OMSC group and compared to those of the benchmark group, effectively comparing what the equivalent bed days and costs per-patient-readmitted would be from a group the same size as the OMSC group but with readmission rates, bed days and costs of the benchmark group.

## 3 Results

Six hundred and seventy three patients who smoked were admitted to hospital and assessed by a TDS between July 2020 and June 2021. Of the 299 who were successfully contacted at six months after discharge from hospital, 104 (34.8%) reported being a non-smoker and 195 (65.2%) reported still smoking. A further 37 patients (5.5%) had opted out of the intervention prior to the six month mark. The sociodemographic and clinical characteristics of the cohort are shown in **Supplementary Table S2**.

### 3.1 Cost per quit

The total cost of the adapted OMSC program between July 2020 and June 2021 was £178,105. This encompassed £130,061 staffing costs for three full-time equivalent TDSs and 0.1 full-time equivalent respiratory medical consultant, £15,893 NRT and pharmacotherapy costs (within 200 days of patient discharge), £21,637 pharmacy staff costs, and £10,512 Trust overhead costs. Dividing by the 673 patients in the OMSC group results in a cost per patient of £264.64, and dividing by the 104 patients who reported being a non-smoker at six months after discharge results in a cost per quit of £1712.55. Using the tables and formula provided in Stapleton & West (2012), the age-adjusted incremental cost per life year gained was £3325 (see **Supplementary Material S3** for details of calculation).

### 3.2 Benchmark analysis

After excluding patients who died within the year following discharge (n=22), patients with missing follow-up data (n=20), and patients who could not be benchmarked (n=20), 611 patients from the OMSC group were included in the final benchmark analysis (mean age = 50.6 years, SD = 16.1). Of the OMSC group, 5.2% (n=32) were readmitted after six months from their discharge date. The benchmark cohorts consisted of 7312 matched patients in total (mean age = 49.4, SD = 18.2), of whom 11.1% were readmitted in the same timeframe (n=812).

The total readmission costs of the OMSC group were £266,288, and the averaged readmission costs from the benchmark group (n=611) were £757,811. This represents a total saving of £491,523 associated with the OMSC intervention. The OMSC group also incurred 414 fewer readmission bed days than the benchmark group (303 vs 717 bed days). In terms of per-patient-readmitted differences, the OMSC group had 1.2 fewer bed days and cost £2904.76 less per-patient-readmitted. See **Table 1** for comparison of total readmission rates, costs, and bed days per patient readmitted for the OMSC group and benchmark group.

**Table 1.**
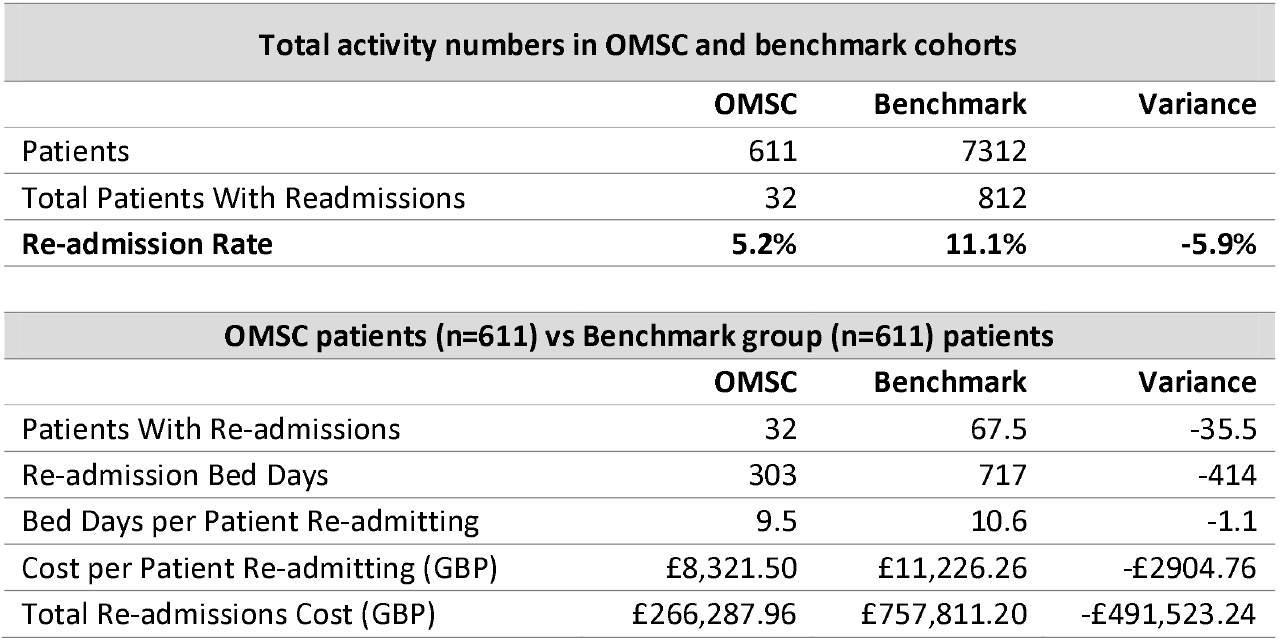
Comparison of total readmission rates, costs, and bed days per patient readmitted, for the OMSC group and the equivalently-sized benchmark group.

| Total activity numbers in OMSC and benchmark cohorts |  |  |  |
| --- | --- | --- | --- |
|  | OMSC | Benchmark | Variance |
| Patients | 611 | 7312 |  |
| Total Patients With Readmissions | 32 | 812 |  |
| **Re-admission Rate** | **5.2%** | **11.1%** | **-5.9%** |

| OMSC patients (n=611) vs Benchmark group (n=611) patients |  |  |  |
| --- | --- | --- | --- |
|  | OMSC | Benchmark | Variance |
| Patients With Re-admissions | 32 | 67.5 | -35.5 |
| Re-admission Bed Days | 303 | 717 | -414 |
| Bed Days per Patient Re-admitting | 9.5 | 10.6 | -1.1 |
| Cost per Patient Re-admitting (GBP) | £8,321.50 | £11,226.26 | -£2904.76 |
| Total Re-admissions Cost (GBP) | £266,287.96 | £757,811.20 | -£491,523.24 |

#### 3.2.1 Stratification by 6-month quit status

See **Figures 2** and **3,** and **Supplementary Table S4**, for comparisons of the OMSC and benchmark groups in terms of readmission rates, total readmission costs and bed days, and mean costs and bed days per-patient-readmitted, according to OMSC group smoking status at six months after discharge.

**Figure 2.**
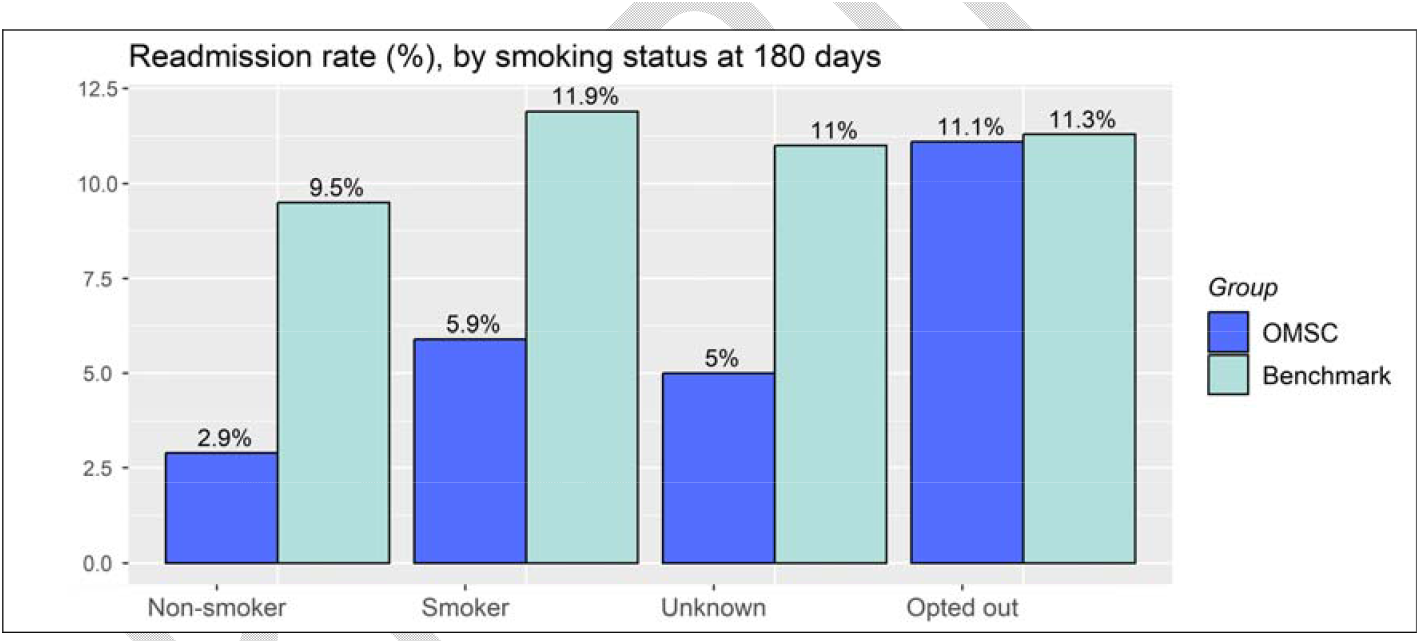
Readmission rates compared between the OMSC group (n=611) and their benchmark group equivalents (n=611), according to OMSC group smoking status at six months after discharge.

**Figure 3.**
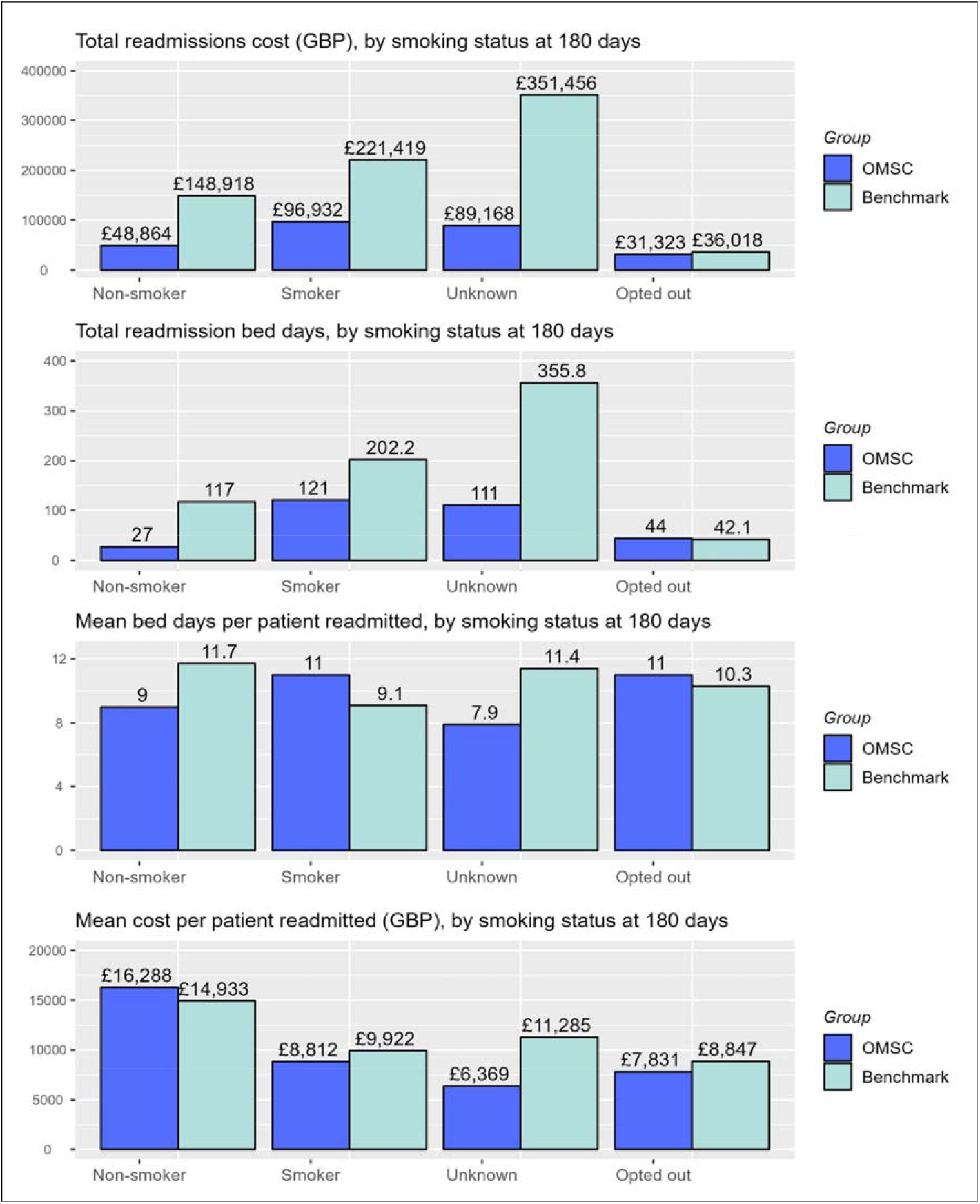
Comparisons between the OMSC group (n=611) and their benchmark group equivalents (n=611), according to OMSC group smoking status at six months after discharge: total readmissions cost (GBP), total readmission bed days, mean bed days per-patient

When stratified according to OMSC group smoking status at six months, lower readmission rates and correspondingly lower costs were found among all smoking status categories for the OMSC group when compared to their benchmark group equivalents, except among those who opted out of the OMSC intervention for whom there was no difference. Compared to their matched benchmarks, mean bed days per-patient-readmitted were lower for the non-smoker and unknown smoking status OMSC subgroups, and marginally higher for the smoker and opt-out groups. Despite the cost per-patient-readmitted being highest for the non-smokers in the OMSC group and their benchmark equivalents, due to the difference in readmission rates between OMSC and benchmark groups the OMSC group data still reveals significant cost savings. Of note, the mean age of the readmitted OMSC patients was highest in the non-smoking group (64.3 years [benchmark = 61.7]) compared to readmitted patients in the smoking (53.9 years [benchmark = 55.9]), unknown (54.0 years [benchmark = 51.9]), and opt-out (57.0 years [benchmark = 53.0]) groups.

## 4 Discussion

### 4.1 Summary of findings

An inpatient smoking cessation intervention based on the OMSC was implemented for one year in an acute hospital setting in London. Among 673 patients who were seen by the TDS, of the 299 who were able to be contacted six months after discharge, 104 reported being a non-smoker. The cost per quit was estimated to be £1713, with an age-adjusted ICER of £3325 per life year gained. Among the 611 patients who were able to be matched to a benchmark cohort of similar patients who did not receive the OMSC intervention, re-admissions for patients in the OMSC group cost £492k less than their benchmark equivalents for the 21 months from January 2021 to September 2022 (£266k vs £758k), incurred 414 fewer bed days (303 vs 717), and re-admitted at a lower rate (5% vs 11%).

Applying the assumption used by the NHS—that approximately 50% of the fiscal benefits can be cashable if the scale of the transformation programme is large (13)—gives a fiscal benefit of £245,762 for the 21 months from January 2021 to September 2022, against the cost incurred for the intervention of £178,105 during the 12-month period from July 2020 to June 2021. This equates to a return on investment of £1.37 for every £1 spent. Considering this was a pilot intervention implemented during the unprecedented disruption of the first year of the COVID-19 pandemic, in which over half of the admitted smokers were not seen by a TDS, suggests a potential for greater returns on investment following expansion of the intervention, with public health interventions typically generating greater cashable cost savings over longer periods of time and when implemented at scale (17). However, the costs of the OMSC intervention established here already place it well below the threshold of cost-effectiveness used by the National Institute of Health and Care Excellence (NICE) in England; ICERs of less than £20,000 per QALY gained are considered cost-effective (18). Furthermore, the financial benefits of quitting smoking accrue not only to the NHS but to the individual patients who quit, their families, and wider society through increases in healthy life expectancy (2), and reductions in health inequalities (19). In the London borough in which King’s College Hospital in situated, an estimated £85.6M is spent on tobacco, costing the borough £206M per year, with £11.2M spent of healthcare costs alone (20).

Direct comparisons to other studies are difficult, due to variation in settings, type of patient, length of follow-up and modelling techniques (12). The closest comparison in terms of setting comes from the CURE Project pilot in Manchester, UK, which reported a lower cost per quit of £475 and ICER of £487 per QALY gained, however this was based on quit rates at 12-weeks rather than 6-months after discharge, and a cohort of more than double the size (n=1450) (13). By contrast, a Dutch trial of an inpatient smoking cessation intervention for patients with coronary heart disease, followed up for 6-months, found approximately £4100 (€4781) of healthcare costs per quit, based on a group of 157 patients receiving face-to-face counselling (21).

In our benchmarking analysis, the beneficial variance in readmission costs and bed days was primarily driven by reduced readmission rates in the OMSC group. Whilst the greatest variance was found among those who reported not smoking a six-month follow-up (2.9% vs 9.5% benchmark), similar reductions in readmission rate were found among those OMSC patients whose six-month smoking status was unknown (5% vs 11% benchmark) and among those who reported still smoking (5.9% vs 11.9%). The only group for whom no meaningful reduction in readmission rate occurred were those patients who opted out of the OMSC intervention (11.1% vs 11.3% benchmark). Whilst benefits derived in the unknown smoking status group might be due to some patients quitting smoking but not answering the phone to confirm, the benefits derived among the reported still-smoking group are more challenging to interpret, and run counter to oft-used principle that no health benefit attributable to the intervention is assumed for those still smoking at final follow-up (22). However, it is possible that simply being exposed to the intervention prompts some future behaviour change resulting in reduced hospital readmission; for example, a trial of motivational tobacco cessation treatment combined with NRT in psychiatric inpatients found that being in the treatment group was associated with significantly decreased odds of rehospitalisation within 18 months compared to the usual-care control group, but actual quit success was not (23). It is also possible that the use of a point-prevalence measure of smoking status at six months conceals some quit success in the months leading up to that point, wherein a temporary relapse may have occurred.

A further unanticipated finding was that the mean costs per-patient-readmitted were highest among the OMSC patients who reported not smoking at six-month follow-up and their benchmarks, compared to all other smoking status groups. However, this might be explained by the readmitted non-smokers being a particularly small group (n<10)^1^ with an older mean age (64 years) compared to readmissions in other smoking status categories (mean=55 years).

### 4.2 Strengths and Limitations

Our study uses a well-validated PLICS tool to attribute accurate healthcare costs to patient activity; an assessment of the Trust’s costing tool’s data quality and adherence to NHS Costing Transformation Programme standards was scored at 98% and has been ranked in the top 5 nationally (24). In terms of limitations, the estimated costs do not include wider societal costs, and do not include post-discharge costs such as local authority Stop Smoking Services, to which some patients were referred.

We do not know the post-discharge smoking status of any of the benchmark patients, as they were not part of the TDS’s caseload, and so stratification according to 6-month smoking status is based purely on OMSC group smoking status, and smoking status was not biochemically verified. Delayed transfer of care could confound the readmission bed day calculations, although this risk applies equally across OMSC and benchmark groups, and so should not be a significant source of bias.

### 4.3 Conclusions

Our economic evaluation of an adapted Ottawa model of smoking cessation intervention implemented in a major acute London hospital found the intervention to be sufficiently cost-effective. Reduced readmission rates and consequent costs were found for patients who received the intervention compared to their benchmarked equivalents, except for those who opted out.

## Data Availability

This study uses de-identified patient data and as such cannot be made available. For queries, please contact the corresponding author

## Footnotes

1 Frequencies fewer than 10 supressed to prevent potential disclosure of patient-identifiable information (25)

