## Supplementary material for "Economic evaluation of a hospital-initiated tobacco dependence treatment service"

S1: Matching rules and loops

|  | Matching Loop Order Number | | | | | | | | | | | | | |
| --- | --- | --- | --- | --- | --- | --- | --- | --- | --- | --- | --- | --- | --- | --- |
| Matching rule | **1** | **2** | **3** | **4** | **5** | **6** | **7** | **8** | **9** | **10** | **11** | **12** | **13** | **14** |
| Smoker | ✓ | ✓ | ✓ | ✓ | ✓ | ✓ | ✓ |  |  |  |  |  |  |  |
| How Many Smoked | ✓ | ✓ | ✓ | ✓ | ✓ | ✓ | ✓ | ✓ | ✓ | ✓ | ✓ | ✓ | ✓ | ✓ |
| Point of Delivery | ✓ | ✓ | ✓ | ✓ | ✓ | ✓ | ✓ | ✓ | ✓ | ✓ | ✓ | ✓ | ✓ | ✓ |
| HRG Root | ✓ | ✓ | ✓ | ✓ |  |  |  | ✓ | ✓ | ✓ | ✓ |  |  |  |
| HRG Sub-Chapter |  |  |  |  | ✓ | ✓ | ✓ |  |  |  |  | ✓ | ✓ | ✓ |
| IMD Decile | ✓ | ✓ |  |  | ✓ | ✓ |  | ✓ | ✓ |  |  | ✓ | ✓ |  |
| Age Range | ✓ |  | ✓ |  | ✓ |  | ✓ | ✓ |  | ✓ |  | ✓ |  | ✓ |
| Min. 5 Spells in Bench cohort | ✓ | ✓ | ✓ | ✓ | ✓ | ✓ | ✓ | ✓ | ✓ | ✓ | ✓ | ✓ | ✓ | ✓ |
| % Patients Matched | **7%** | **26%** | **7%** | **28%** | **1%** | **8%** | **3%** | **0%** | **1%** | **0%** | **10%** | **0%** | **2%** | **7%** |
| Count of Patients Matched | **45** | **158** | **44** | **169** | **6** | **47** | **19** | **0** | **8** | **0** | **59** | **0** | **15** | **41** |

***Supplementary Table S1:***

*The matching rules applied to patient records, used to identify benchmark cohorts matched to the OMSC group patients*

*HRG = Healthcare Resource Group (clinically meaningful groupings of patient activity derived from NHS patient records, primarily using procedure and diagnosis codes, see* [*www.digital.nhs.uk/services/secondary-uses-service-sus/payment-by-results-guidance/sus-pbr-reference-manual/hrg-grouping*](http://www.digital.nhs.uk/services/secondary-uses-service-sus/payment-by-results-guidance/sus-pbr-reference-manual/hrg-grouping)*); IMD = Index of Multiple Deprivation*

S2: Characteristics of OMSC group cohort

| **Variable** | **Category** | **n (%)** |
| --- | --- | --- |
| **Age** | 60+ | 228 (33.9%) |
|  | 40-59 | 260 (38.6%) |
|  | 25-39 | 144 (21.4%) |
|  | 16-24 | 41 (6.1%) |
| **Sex** | Male | 422 (62.7%) |
|  | Female | 251 (37.3%) |
| **Ethnicity** | Asian | 13 (1.9%) |
|  | Black | 125 (18.6%) |
|  | Mixed | 21 (3.1%) |
|  | Other | 63 (9.4%) |
|  | White | 389 (57.8%) |
|  | Declined or Not stated | 62 (9.2%) |
| **HSI category** | Low | 194 (28.8%) |
|  | Medium | 294 (43.7%) |
|  | High | 42 (6.2%) |
|  | *Missing* | *143 (21.2%)* |
| **Primary diagnosis** | Circulatory | 54 (8.0%) |
|  | Digestive | 99 (14.7%) |
|  | Endocrine and blood | 39 (5.8%) |
|  | Genitourinary | 31 (4.6%) |
|  | Infectious and parasitic | 26 (3.9%) |
|  | Injury poisoning and external | 179 (26.6%) |
|  | Mental Behavioural & Neurodevelopmental | 24 (3.6%) |
|  | Musculoskeletal | 20 (3.0%) |
|  | Neoplasms | 39 (5.8%) |
|  | Nervous system | 15 (2.2%) |
|  | Respiratory | 60 (8.9%) |
|  | Skin and subcutaneous | 12 (1.8%) |
|  | Other | 29 (4.3%) |
|  | *Missing* | *46 (6.8%)* |
| **IMD tertile** | Lower | 329 (48.9%) |
|  | Middle | 193 (28.7%) |
|  | Upper | 59 (8.8%) |
|  | *Missing* | *92 (13.7%)* |
| **Six-month smoking status** | Non-Smoker | 104 (15.5%) |
|  | Smoker | 195 (29.0%) |
|  | Unknown | 295 (43.8%) |
|  | Opted out | 37 (5.5%) |
|  | Patient deceased | 22 (3.3%) |
|  | *Missing* | *20 (3.0%)* |

**Supplementary Table S2**: Characteristics of cohort (N=673).

*IMD = Index of Multiple Deprivation, HSI = Heaviness of Smoking Index, Primary diagnoses grouped according to ICD-10 chapter*

### S3: Incremental Cost-Effectiveness Ratio

Incremental Cost-Effectiveness Ratio = the ratio of the increased cost per person to provide the intervention (incremental cost, IC) and the health benefit per person (incremental effectiveness, IE): ICER = IC/IE.

**Intervention effect** =

- 104 non-smokers at 6 months / 673 in cohort = 15.5%
- Minus 2.5% background quit rate in general population = 13%

**Intervention cost** = £264.64 per smoker treated

**ICER interpolated from Table 3A in (Stapleton & West, 2012)** = £2634

**Adjustment for age** **distribution in OMSC group** (see Stapleton & West, 2012) =

- Over 54 = 47.1% (multiply ICER by 1.36)
- 35-54 = 32.7% (ICER as above)
- Under 35 = 20.2% (multiply ICER by 1.46)

**Put it all together =**

- 0.202 * (1.46 * 2634) + 0.327 * 2634 + 0.471 * (1.36 * 2634) = **£3325.37**

***So this would mean the OMSC intervention costs £1712.55 per quit at 6 months, and £3325 per Life Year gained.***

**Assumptions:**

- assumes that there is no benefit to anything other than permanent cessation, which is unlikely to be true
- assumes that all of the 295 patients whose smoking status was unknown at six months follow-up continued to smoke
- assumes 3.5% discounting rate (i.e. later years = less value)
- assumes that 2.5% of smokers would have quit anyway, regardless of intervention
- assumes 48.75% of those abstinent at 6 months will remain so (others relapse)

**See:**

Stapleton, J. A., & West, R. (2012). A Direct Method and ICER Tables for the Estimation of the Cost-Effectiveness of Smoking Cessation Interventions in General Populations: Application to a New Cytisine Trial and Other Examples. *Nicotine & Tobacco Research*, 14(4), 463–471. <https://doi.org/10.1093/ntr/ntr236>

S4: Variance in costs and bed days between OMSC and benchmark groups, stratified by smoking status at 6 months

| **OMSC group 6-month smoking status** | **Group** | **Readmission rate** | **Total readmissions cost** | **Total readmission bed days** | **Mean cost per-patient-readmitted** | **Mean bed-days per-patient-readmitted** |
| --- | --- | --- | --- | --- | --- | --- |
| **Non-smoker** | OMSC | 2.9% | £48,864.18 | 27.0 | £16,288.06 | 9.0 |
| n=104 | Benchmark | 9.5% | £148,917.70 | 117.0 | £14,933.48 | 11.7 |
|  | *Variance* | *-6.6%* | *-£100,053.52* | *-90.0* | *£1,354.58* | *-2.7* |
| **Smoker** | OMSC | 5.9% | £96,931.90 | 121.0 | £8,811.99 | 11.0 |
| n=188 | Benchmark | 11.9% | £221,419.20 | 202.2 | £9,921.63 | 9.1 |
|  | *Variance* | *-6.0%* | *-£124,487.30* | *-81.2* | *-£1,109.64* | *1.9* |
| **Unknown** | OMSC | 5.0% | £89,168.41 | 111.0 | £6,369.17 | 7.9 |
| n=282 | Benchmark | 11.0% | £351,456.20 | 355.8 | £11,285.20 | 11.4 |
|  | *Variance* | *-6.1%* | *-£262,287.79* | *-244.8* | *-£4,916.03* | *-3.5* |
| **Opted-out** | OMSC | 11.1% | £31,323.47 | 44.0 | £7,830.87 | 11.0 |
| n=37 | Benchmark | 11.3% | £36,018.08 | 42.1 | £8,846.55 | 10.3 |
|  | *Variance* | *-0.2%* | *-£4,694.61* | *1.9* | *-£1,015.68* | *0.7* |
| **ALL** | OMSC | 5.2% | £266,288.00 | 303.0 | £8,321.50 | 9.5 |
| N=611 | Benchmark | 11.1% | £757,811.20 | 717.0 | £11,226.26 | 10.6 |
|  | *Variance* | *-5.9%* | *-£491,523.20* | *-414.0* | *-£2,904.76* | *-1.2* |

***Supplementary Table S4*** *Readmission rates, total readmissions costs & bed-days, and mean costs & bed days per-patient-readmitted, stratified according to OMSC group smoking status at 6-month follow-up*
